# Comparative Analysis of Muscle Fascia Tracking Algorithms for Real-Time Muscle Monitoring using Wearable Ultrasound

**DOI:** 10.64898/2026.07.30.26359276

**Authors:** Erica L. King, Connor Delaney, Morgan A. Lamarre, Amad Qureshi, Siddhartha Sikdar, Qi Wei, Parag V. Chitnis

## Abstract

Musculoskeletal ultrasound (MSK-US) enables real-time imaging of muscle structure and function, and wearable ultrasound (WUS) has extended this capability to dynamic movement tasks. Accurate tracking of muscle fascia displacement in M-mode WUS images is essential for quantifying muscle function, yet the relative performance of existing fascia-tracking algorithms remains uncharacterized. This study directly compares five fascia-tracking algorithms: Maximum Pixel Intensity (MPI), Muscle Boundary Tracking Algorithm (MBTA), Principal Component Analysis (PCA), Composite-Factorization PCA (CF-PCA), and U-Net segmentation, against expert-annotated ground truth to identify which approach best supports wearable muscle-monitoring applications. A total of 572 M-mode ultrasound images were collected during isometric quadricep activations (QA) and squats (SQ) using a multi-site WUS system with transducers positioned on the vastus lateralis (VL), rectus femoris (RF), and vastus medialis oblique (VMO). Fascia tracking using U-Net segmentation exhibited the lowest mean absolute error (median QA=0.57, median SQ=1.22; p<0.05), functional range not statistically different from expert traces (QA p=0.33; SQ p=1) and the most accurate estimates of functional error (median QA=-0.21; median SQ=-0.65; p<0.05). PCA-based methods demonstrated the highest correlation with the expert traces (PCA median QA=0.88; CF-PCA median QA=0.88; PCA median SQ=0.78; CF-PCA median SQ=0.75; p<0.005), reflecting superior tracking of relative contraction patterns. These results indicate U-Net segmentation is best suited for applications requiring precise fascia-depth estimation when labeled training data are available, while PCA-based methods are preferable for tracking relative contraction patterns without supervised training, informing algorithm selection for wearable neuromuscular monitoring in clinical and performance settings.

## INTRODUCTION

Musculoskeletal ultrasound (MSK-US) has become a valuable tool for real-time imaging of muscle structure and function [1]. MSK-US imaging has shown the ability to image and characterize musculoskeletal tissues such as muscle compartments, tendons, and soft tissues to determine the severity of injury [2], [3], [4]. Despite the promise of MSK-US, quantitative outcome measures are hampered by several practical challenges. Even with recent, rapid MSK-US advancements and the introduction of point-of-care ultrasound and wireless, handheld ultrasound systems, transducers continue to be designed for handheld operation, with significant operator dependence in acquiring and interpreting MSK-US images [5], [6].

To address this, wearable ultrasound (WUS) systems have emerged, enabling real-time assessment of muscle function during movement [7]. Current applications of WUS primarily focus on cardiovascular and pulmonary monitoring, as well as exoskeleton and prosthesis control [8], [9], [10], [11], [12], [13]. Beyond these areas, WUS offers tremendous potential for simultaneously imaging multiple muscle groups involved in force production during movement, enabling a more comprehensive and systematic assessment of muscle function [14]. The evaluation of muscle groups, including muscles which are not superficial, during rehabilitation training following injuries or in individuals with motor disorders is a unique strength of WUS as contributions from separate muscles involved in the same task can be captured. This includes those under the same transducer. Moreover, stroke rehabilitation and strength training in young, able-bodied individuals incorporating WUS-based biofeedback has been shown to improve muscle force generation and contractility [15], [16].

The intended end user for this technology is not uniform. Care providers without ultrasound-specific training are unlikely to incorporate MSK-US into clinical decisions without substantial upskilling, while neuromuscular specialists and sports medicine clinicians who already read ultrasound competently currently lack a standardized, quantitative output they can track across sessions or compare against a patient’s baseline. Moreover, there are still many refinements required to push WUS systems from research devices to clinical tools that serves both groups of clincians, spanning the imaging methods themselves and the automated, standardized outputs derived from them.

One such refinement is the development of imaging techniques which maintain utility but are also lightweight and scalable across multiple recording sites. This has led researchers to explore motion-mode (M-mode) ultrasound as an alternative to traditional brightness-mode (B-mode) techniques for capturing time-varying muscle deformation [17]. Previous studies have demonstrated that M-mode ultrasound imaging can serve as a practical alternative to B-mode imaging for investigating time-varying muscle deformation [14], [18]. M-mode captures ultrasound scanlines over time from a single fixed position, allowing muscle motion to be monitored more efficiently [19], [20]. This approach simplifies system design by reducing electronic complexity, minimizing device footprint, lowering computational demands, and facilitating the integration of multiple imaging sensors capable of monitoring multiple sites.

Accurate tracking of the displacement of muscle fascia within M-mode images are useful for quantifying characteristics of muscle function through contraction timing, changes in muscle thickness, and bilateral symmetry [21], [22], [23]. These measures may be used by clinicians to quantify muscle behavior in patients or incorporated as signals presented to users in training using biofeedback, supporting the broader case for ultrasound-derived measures as an outcome measure in rehabilitation and performance settings rather than solely a diagnostic tool [24]. Historically, fascia tracking has been performed manually by experts, a process that is time-consuming and prone to subjective bias [25], [26], [27]. As WUS systems become more prevalent, especially in non-laboratory environments, there is a growing need for automated fascia tracking algorithms that can operate reliably under real-world dynamic conditions.

Several deterministic tracking algorithms have been proposed to automate fascia detection [22], [28], [29], [30], [31]. The most basic method, Maximum Pixel Intensity (MPI) tracking, identifies the brightest pixel in each frame as the fascia location [30]. While simple and widely used, MPI is highly sensitive to brightness fluctuations caused by probe movement, pressure changes, or gel coupling inconsistencies, leading to inaccurate tracking. To overcome these limitations, Jin et al. developed a hybrid and data-driven approach called the Muscle Boundary Tracking Algorithm (MBTA), which combines MPI with another traditional tracking technique, cross-correlation-based tracking, to balance low-frequency stability and high-frequency sensitivity [29]. MBTA, however, still faces challenges, including sensitivity to probe pressure changes, susceptibility to imaging-plane shifts, and reduced robustness during rapid movements.

To address these issues, Principal Component Analysis (PCA) fascia tracking offers a more robust alternative by capturing dominant patterns of variance in pixel depth vectors, making it less susceptible to probe movement and imaging plane shifts, which stem from decorrelations [28]. Despite these advantages, PCA tracking is limited by its reliance on global variance patterns, which reduces sensitivity to localized fascia motion and subtle interface changes during dynamic contractions. Building on this, we have developed a Composite Factorization PCA (CF-PCA) algorithm, which integrates multiple levels of abstraction from non-negative matrix factorizations to enhance sensitivity to interface motion [27].

In addition to deterministic fascia tracking algorithms, deep learning approaches have been used for image segmentation [32], [33]. U-Net architectures have demonstrated strong performance in biomedical image segmentation tasks due to their ability to learn hierarchical features and localize structures with high accuracy [34]. The encoder-decoder design of U-Nets enables efficient feature extraction at multiple scales, while skip connections preserve spatial detail, making it highly effective for delineating tissue boundaries in both large and small amplitude movements [33], [34]. These characteristics position U-Nets as a promising candidate for automating fascia tracking in M-mode ultrasound, where dynamic movements introduce variability that traditional algorithms struggle to handle. Furthermore, U-Nets have been used to segment M-mode US images of bones and cardiac muscle with success, indicating MSK-US may similarly be segmented successfully with the architecture [33], [35].

Although these advancements represent significant progress, the performance of tracking algorithms often degrades during dynamic movements, where image characteristics change rapidly [22]. Given the wide range of fascia tracking algorithms, a systematic evaluation is essential to determine their effectiveness across different movement tasks. The objective of this study is to conduct such an evaluation by examining both established and original tracking approaches comparing them to expert-annotated M-mode US images collected during isometric quadriceps activations and bodyweight squats. Algorithm outputs are compared to manual traces using measures of error and accuracy to identify strategies that optimize performance, efficiency, and scalability for fascia tracking in WUS applications. These measures include the mean absolute error (MAE) for fascia depth accuracy, Pearson’s correlation representing trace shape reconstruction, functional range (FR) and functional error (FE) describing the contraction range characteristics captured by traces, bilateral symmetry index (BSI) for quantifying the symmetry between right and left sides during movements, and computational time. The error measures are selected to provide a direct evaluation of the tracking performance on each M-mode image while functional measures provide an indication if the clinically relevant characteristics of the movement are properly represented by the traces.

This evaluation provides a practical guide for researchers using M-mode US to monitor muscle fascia movement by clarifying the strengths and limitations of both traditional and newly developed tracking algorithms. Although the scope of this study does not encompass all possible tracking methods or movement types, it establishes a foundational framework for future investigations into muscle contraction dynamics and supports the broader adoption of WUS technologies in both clinical and performance settings. Accordingly, this study presents a systematic comparison of five fascia tracking paradigms evaluated on wearable M-mode ultrasound data using complementary signal-level, functional-level, and computational metrics.

The contributions of this study are threefold:

1. We present the first systematic comparison of five muscle fascia tracking paradigms—intensity-based, correlation-based, variance-based, and deep learning–based—applied to wearable M-mode ultrasound data acquired during both isometric and dynamic movements.
2. We evaluate tracking performance using complementary signal-level and functional-level metrics, enabling assessment of both trace fidelity and the reliability of derived biomechanical measures such as functional range, functional error, and bilateral symmetry.
3. We provide practical guidance for algorithm selection in wearable musculoskeletal sensing applications by characterizing trade-offs among accuracy, robustness, computational cost, and training requirements, with particular relevance to real-time and resource-constrained deployments.

## METHODS

### A. Movement Tasks and MSK-US Data Acquisition

Twenty-five participants (1 male, age 24 years, height 165.1 cm, weight 79.4 kg, BMI 29.1; 24 females, age 21 ± 2 years, height 173.2 ± 8.5 cm, weight 74.8 ± 10.0 kg, BMI 25.1 ± 4.1) performed quadricep activations (QA) and squats (SQ) while wearing a multi-site WUS. Participants completed up to four experimental sessions spaced at least 48 hours apart: 6 participants completed all four sessions, 1 completed two sessions, and 18 completed one session. All participants were cleared for participation, received a full explanation of the study procedures, and provided written informed consent.

Isometric quadriceps activation and bodyweight squats were selected because both are standard components of clinical lower-extremity assessment and rehabilitation protocols, particularly following knee injury (e.g., ACL reconstruction), where quadriceps strength and functional squat performance are primary return-to-activity benchmarks [36]. These tasks also span two distinct contraction types, static isometric loading and dynamic, multi-joint movement, allowing the tracking algorithms to be evaluated under both simple and functionally complex conditions relevant to real-world monitoring.

Three isometric QA trials were performed each experimental session, with participants lying supine. Each trial consisted of five bilateral maximal quadriceps contractions (5 seconds each) with metronome-paced timing and rest between trials. For QA trials, single-element PZT transducers were placed bilaterally on the vastus lateralis (VL) and rectus femoris (RF). Participants also performed six SQ trials at each session, three for each of two transducer configurations. The first configuration was the same as used for QA, with transducers on the VL and RF bilaterally. For the second configuration, transducers were placed bilaterally on the VL and vastus medialis oblique (VMO). These muscle sites were selected based on prior evidence of maximal activation during the targeted movements [14], [37]. Each SQ trial consisted of three bodyweight squats at a self-selected pace and depth. US data from all four transducers were recorded as individual M-mode images for every repetition for algorithm evaluation (Fig. 1C). The study was approved by the George Mason University Institutional Review Board and conducted in accordance with the Declaration of Helsinki.

**Fig. 1.**
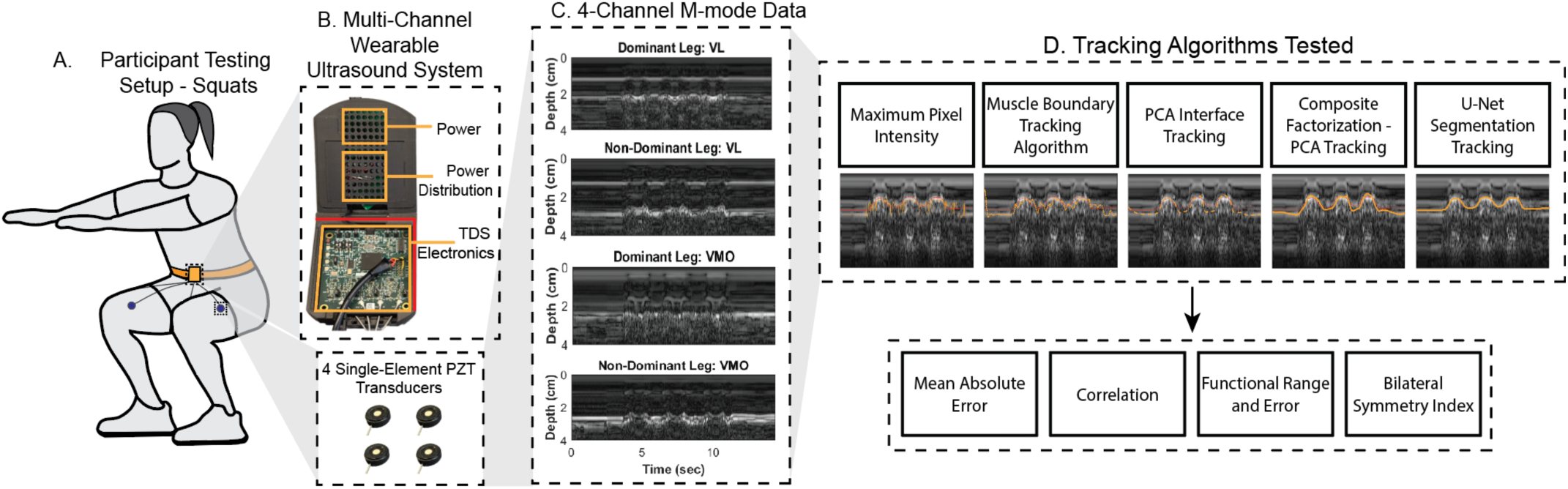
Overview of the experimental setup and analysis pipeline for wearable ultrasound during squats. The same setup and pipeline were completed for isometric quadricep activations. (A) Participant testing setup showing squat exercise with wearable ultrasound transducers placed to image the quadriceps. (B) Multi-channel wearable ultrasound system components, including power distribution, TDS electronics, and single-element PZT transducers. (C) Example 4-channel M-mode ultrasound data collected from dominant and non-dominant legs (VL: vastus lateralis; VMO: vastus medialis oblique) during bilateral body-weight squats over time. (D) Tracking algorithms tested for muscle boundary detection, including Maximum Pixel Intensity, Muscle Boundary Tracking, PCA Interface Tracking, Composite Factorization–PCA Tracking, and U-Net Segmentation Tracking. Performance metrics include Mean Absolute Error, Correlation, Functional Range and Error, and Bilateral Symmetry Index.

The dataset resulting from this experiment included 572 M-mode ultrasound images, comprising 308 isometric quadricep activations (QA) and 264 bodyweight squats (SQ).

### B. Wearable Ultrasound Imaging System and Sensor Placement

A custom multi-site WUS imaging system was used to monitor muscle tissue dynamics during the movement tasks (Figs. 1A and 1B). This system employs a patented time delay spectrometry (TDS) paradigm to generate M-mode US images by transmitting a known frequency sweep (3–5 MHz over 10 ms) and recording the returning echoes simultaneously [38], [39]. The received signals are demodulated and converted into A-lines using a Fourier transform, producing high-resolution temporal profiles of tissue displacement [17].

The WUS system supports simultaneous data acquisition from single-element piezoelectric (PZT) transducers at four anatomical sites at a frame rate of 50 Hz and a depth of 4 cm. The main housing unit contains the signal generator, power supply, analog-to-digital converter, and data transfer components, worn around the waist. Each single-element PZT transducer was housed in a custom 3D-printed housing (Fig. 1B).

To ensure consistent skin contact and minimize motion artifacts, all transducers were secured to the skin using kinesiology therapeutic tape.

### C. Data Preparation – Expert Traces

Each M-mode US image was saved as a grayscale 8-bit image and manually annotated in ImageJ [40], by a single annotator with 2 years of experience in musculoskeletal ultrasound image analysis and formal training in the WUS system’s image acquisition protocol, to identify muscle contraction and fascia displacement during the movement tasks. Annotated traces were converted into binary mask images and served as ground truth for evaluating the tracking algorithms. To prepare the traces for analysis, each mask was skeletonized and smoothed using a Savitzky-Golay filter via MATLAB’s *smoothdata* function with a window size of 10 [41], [42].

### D. Data Preparation – Region of Interest (ROI) Selection

For each M-mode image, an ROI was defined around the expert-traced muscle interface with a 2.5 mm margin above and below its extreme positions. This standard approach isolates relevant features for analysis while minimizing interference from unrelated anatomical structures. The preprocessed ROI served as input for all tracking algorithms, with the expert-traced interface depth as ground truth for performance evaluation, and the same ROI was applied consistently across algorithms to ensure a fair comparison of tracking performance. Because the same expert-defined ROI was applied uniformly across all algorithms and all images, ROI selection does not advantage any specific tracking method and does not affect relative performance comparisons reported in this study. Automated ROI selection was not investigated in this study and is acknowledged as an important direction for future work, particularly for real-time and fully autonomous wearable ultrasound applications.

### E. Tracking Algorithms

Five tracking methods were applied to the same ROI: four deterministic algorithms and one deep learning approach (Table I and Fig. 1D).

Maximum Pixel Intensity (MPI) tracking identifies the fascia by selecting the brightest pixel in each frame within the ROI, assuming the interface corresponds to maximum intensity. Restricting the search to the ROI minimizes interference from other bright structures.

The Muscle Boundary Tracking Algorithm (MBTA) combines MPI and cross-correlation to leverage low-frequency stability from MPI and high-frequency sensitivity from cross-correlation. This fusion reduces noise and drift, improving performance in dynamic conditions.

Principal Component Analysis (PCA) extracts the dominant variance source in the image, which may track fascia, assuming displacement drives variance. The tracking technique in the current work differs from previous approaches, as depth vectors across the entire image are used as the input features to the PCA rather than individual scan lines directly [28]. This tracking technique assumes that fascia displacement is the largest contributor to image variance and is therefore captured by the first principal component produced by the PCA. This component is rescaled using bright pixel intensities (i.e., pixels within 10% of the brightest pixel in the ROI) to align with the image depth, then fit with a spline curve to represent interface motion. A final step ensures directional alignment with expert traces by flipping the output if the correlation is below a threshold.

Composite Factorization PCA (CF-PCA) extends PCA-based fascia tracking by incorporating time coefficients from multiple non-negative matrix factorization (NNMF) decompositions to capture multi-scale dynamics. The final component is processed similarly to PCA for fascia displacement estimation.

U-Net segmentation-based tracking was performed using a four-layered model to extract the position of the fascia in each frame of the M-mode image in a pixel-wise fashion from annotated ground truth. ROIs per exercise were analyzed to identify the largest image size, then padded to the nearest multiple of 32 for architecture compatibility and applied across the dataset, before a 70/30 training/testing split (Table II). The split was performed at the image level rather than the subject level, such that images from the same participant possibly appear in both the training and test sets. Furthermore, no data augmentation was performed. Accordingly, U-Net performance reported in this study reflects within-task segmentation capability under consistent imaging conditions, rather than subject-independent generalization. The model was implemented in TensorFlow, and the training parameters used are presented in Table II. Segmentation masks were cropped, skeletonized to a single-pixel wide trace, and smoothed with a Savitzky–Golay filter to produce the final displacement estimate [41].

**TABLE II.**
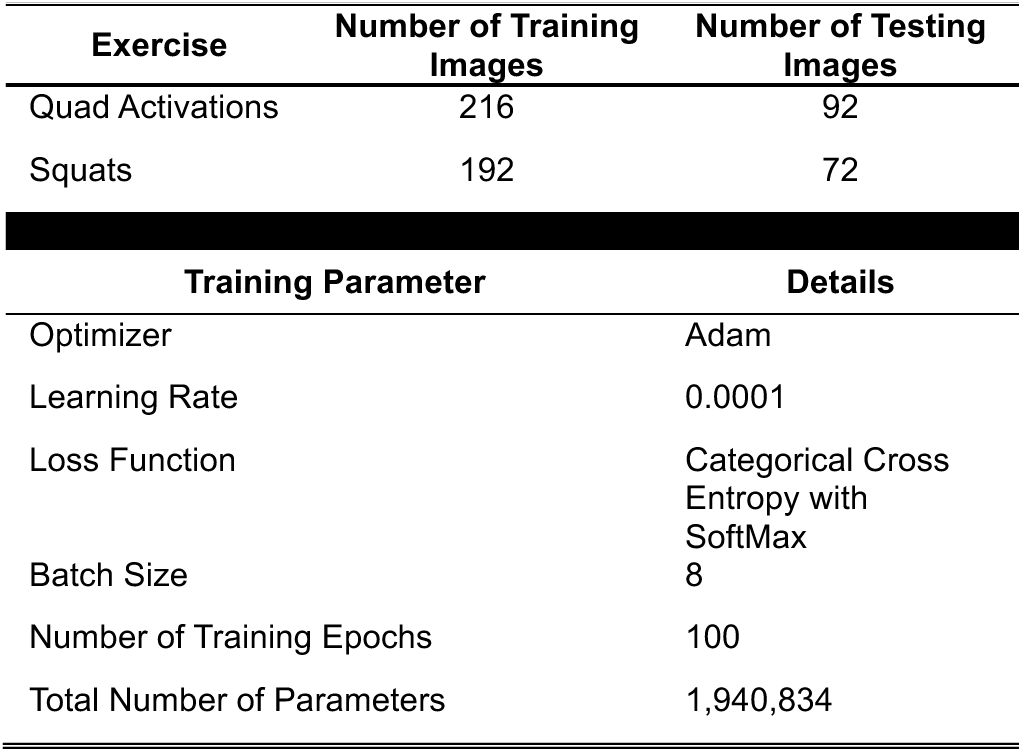
Description of testing and training datasets and training parameters for U-net segmentation tracking.

### F. Statistical Analysis

Algorithm performance was evaluated using five metrics: Mean Absolute Error (MAE), Pearson’s correlation coefficient, Functional Range (FR), Functional Error (FE), and Bilateral Symmetry Index (BSI). MAE quantified pixel-level accuracy between algorithm outputs and expert traces, which provides a direct measure of error in detected fascia depth. Pearson’s correlation assessed similarity in trace shape between algorithms and expert, providing an indication if tracing followed the movements smoothly. FR was calculated as the difference between the mean of the 30 deepest and 30 shallowest points in each trace, representing estimated maximal displacement. FE was defined as the difference between each algorithm’s FR and that of the expert trace. FR provides an indication of the amplitude of contraction is properly captured by tracking algorithms, while FE provides a measure of the error in this metric. Where applicable, BSI was computed to assess symmetry in fascia displacement between dominant leg (DL) and nondominant leg (NDL) muscle groups during the same movement using (1).

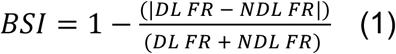

The BSI from all algorithms was compared to the BSI calculated using the expert’s trace to determine if the tracking was capable of being used to calculate a functional measure in the same manner as manual traces from an expert.

To assess the statistical significance of differences across combinations of preprocessing techniques, which are discussed in the Appendix, and tracking algorithms, Kruskal–Wallis tests were applied to all performance metrics except BSI. Non-normality assumptions were verified using Lilliefors tests across all evaluated groups. For BSI, Mann–Whitney U tests compared values derived from expert traces and algorithm outputs. Statistical significance was set at p < 0.05, and Bonferroni correction was applied for multiple comparisons.

In addition to accuracy metrics, computational time was recorded for each algorithm. MPI, MBTA, PCA, and CF-PCA were implemented on a Dell Optiplex 7060 (Intel Core i7-8700 CPU). U-Net segmentation was implemented, and model training was performed on a Lambda Vector workstation with dual NVIDIA RTX A5000 GPUs and CPU fallback (Intel Core i9-10980XE). U-Net training required approximately 360 seconds for QA images and 233 seconds for SQ images.

## RESULTS

### A. Tracking Algorithm Performance – Mean Absolute Error

For both movement tasks, U-Net segmentation had a smaller MAE than the other tracking algorithms (median QA=0.57mm, median SQ=1.22mm; all algorithms on QA p<0.005; PCA and CF-PCA on SQ p<0.05; MPI and MBTA on SQ p<0.005). CF-PCA had significantly smaller MAEs than MPI and MBTA for both movement tasks (QA p<0.05; SQ p<0.005), while PCA had lower MAEs than MPI and MBTA for SQ images only (p<0.005) (Fig. 2 Top row).

**Fig. 2.**
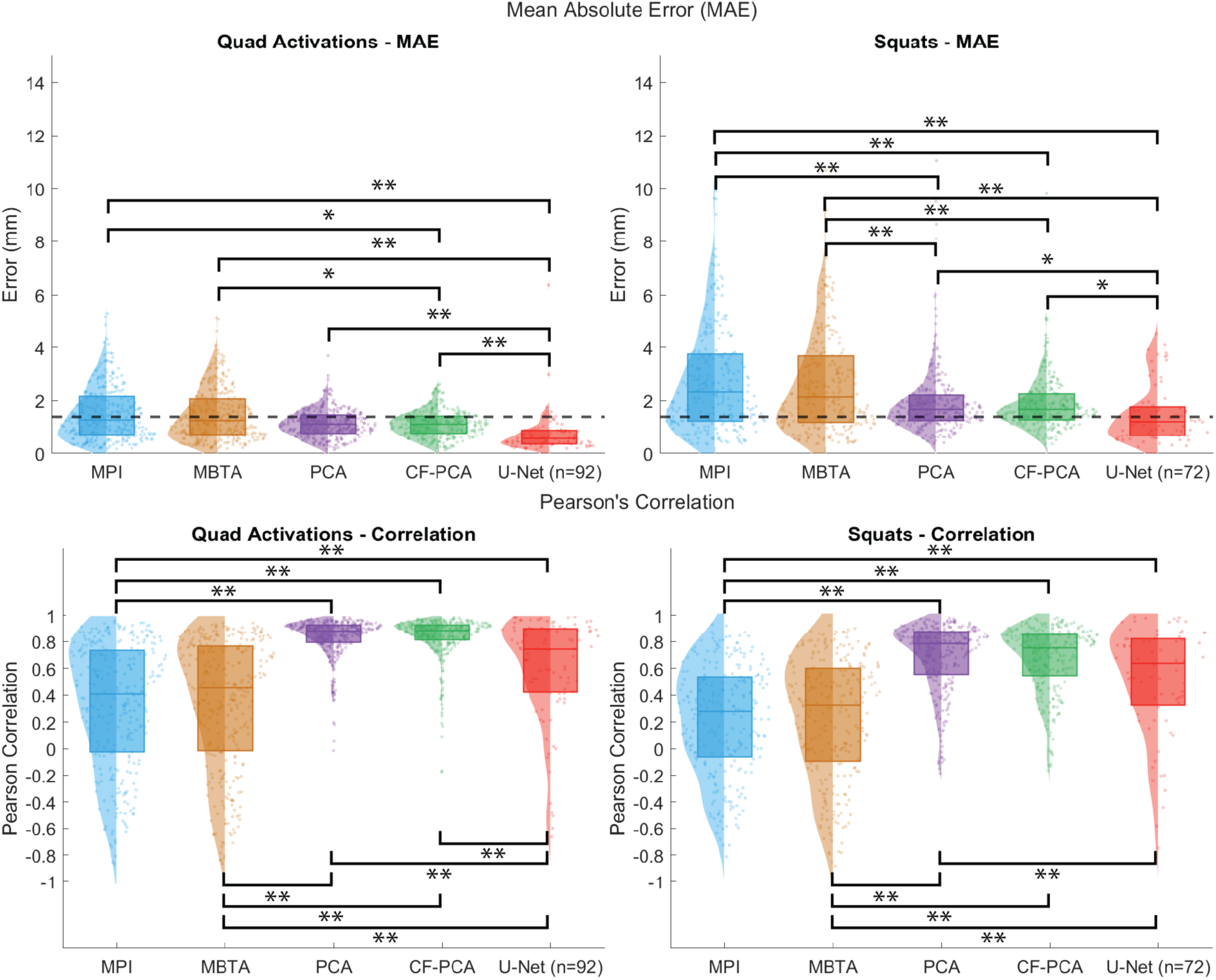
(Top row) Distribution of mean absolute errors (MAE) for the five tracking algorithms applied to QA and SQ. The MAE for each of the tracking algorithms were compared to the original, 10-pixel width (1.4 mm) of the expert traces, shown as the black dashed line. (Bottom row) Distribution of Pearson’s correlation coefficients for the five tracking algorithms applied to QA and SQ. * *p*<0.05 and ** *p*<0.005.

All tracking algorithms applied to the QA images had a median MAE smaller than the thickness of the expert trace (indicated by the dashed line in Fig. 2 Top row). For the SQ images, only the U-Net segmentation had a median MAE smaller than the original width of the expert trace.

### B. Tracking Algorithm Performance - Correlation

MPI and MBTA exhibited significantly lower Pearson’s correlation coefficients than all other tracking algorithms, as well as more variability in their correlations for both movement tasks (p<0.005). PCA had significantly higher correlations than U-Net segmentation for both movement tasks, and CF-PCA had significantly higher correlations than U-Net segmentation when applied to QA images (PCA median QA=0.88; CF-PCA median QA=0.88; PCA median SQ=0.78; CF-PCA median SQ=0.75; p<0.005) (Fig. 2 Bottom row).

### C. Tracking Algorithm Performance – Functional Range, Functional Error, and Bilateral Symmetry Index

For QA images, all tracking algorithms except U-Net segmentation had significantly different FR from the expert trace (p<0.005). However, for SQ images, PCA and CF-PCA FR significantly differed from expert trace FR (p<0.005), while MPI, MBTA, and U-Net segmentation did not (Fig. 3 Top row).

**Fig. 3.**
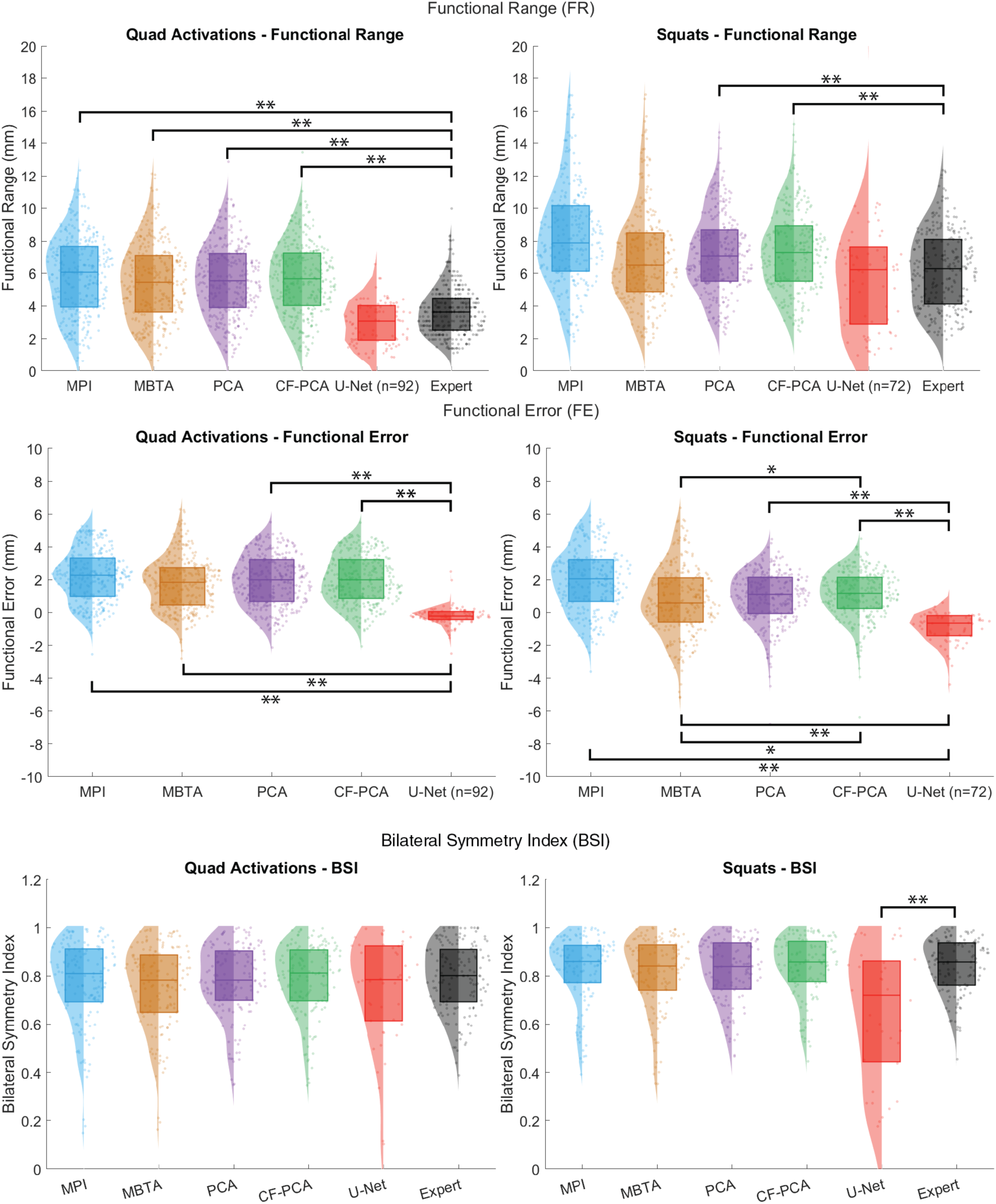
(Top row) Distribution of functional ranges (FR) for the five tracking algorithms applied to QA and SQ. (Middle row) Distribution of functional errors (FE) for the five tracking algorithms applied to QA and SQ. (Bottom row) Distribution of bilateral symmetry indices (BSI) for the five tracking

FE for U-Net segmentation was significantly lower than the other tracking algorithms (median QA=-0.21; median SQ=-0.65; p<0.005). Furthermore, U-Net segmentation had a negative median FE for both movement tasks, while the other tracking algorithms had positive median FE (Fig. 3 Middle row).

All tracking algorithms produced BSI similar to the expert trace BSI with the exception of U-Net segmentation applied to SQ images (p<0.005) (Fig. 3 Bottom row).

### D. Tracking Algorithm Performance – Computational Time

MPI exhibited the fastest computational time (QA 0.002 ± 0.000s; SQ 0.001 ± 0.000s) followed by PCA (QA 0.016 ± 0.005s; SQ 0.008 ± 0.002s) MBTA (QA 0.041 ± 0.002s; SQ 0.022 ± 0.003s), U-Net segmentation (QA 0.104 ± 0.015s; SQ 0.145 ± 0.016s). CF-PCA required the longest computational time out of all the tracking algorithms (QA 7.072 ± 2.560s; SQ 5.724 ± 1.968s). The mean and standard deviation of the time required to produce tracking outputs for the five algorithms are presented in Table III.

**TABLE III.** Mean computational times (sec) for tracking algorithms.

|  | MPI | MBTA | PCA | CF-PCA | U-Net<br>Segmentation | Skeletonization<br>and<br>smoothing |
| --- | --- | --- | --- | --- | --- | --- |
| **QA** | $0.002 \pm 0.000$ | $0.041 \pm 0.002$ | $0.016 \pm 0.005$ | $7.072 \pm 2.56$ | $0.064 \pm 0.007$ | $0.042 \pm 0.008$ |
| **SQ** | $0.001 \pm 0.000$ | $0.022 \pm 0.003$ | $0.008 \pm 0.002$ | $5.724 \pm 1.968$ | $0.125 \pm 0.005$ | $0.020 \pm 0.011$ |

## DISCUSSION

The objective of this study was to characterize and evaluate the relative performance of multiple M-mode ultrasound muscle fascia tracking techniques against one another. The goal of this evaluation was to provide insight into the practical benefits and strengths of each algorithm in a comparative and quantitative manner. Implemented algorithms included both deterministic and deep-learning-based techniques for tracking muscle fascia movement in M-mode US images acquired during dynamic and isometric tasks using a WUS system.

Furthermore, when compared to prior work in muscle US analysis, which has largely focused on B-mode imaging or static tasks, our results seek to underscore the feasibility of dynamic fascia tracking using wearable M-mode systems [43]. By comparing intensity-based methods, PCA-derived approaches, and tracking using a deep learning segmentation model, we identified differences in tracking accuracy and the ability to generate functional measures relevant to clinical and performance applications.

The performance of the five tracking algorithms was assessed by comparing their performance across five metrics using M-mode images from two movement tasks. The U-Net segmentation exhibited the lowest MAE among all the tracking algorithms investigated. The variance-based tracking algorithms (i.e., PCA and CF-PCA) had equivalent errors which were larger than that of the U-net segmentation, yet less than the errors of the intensity-based techniques (i.e., MPI and MBTA). Furthermore, U-Net segmentation performed significantly better than all other algorithms in its ability to obtain approximately the same FR as the expert trace and exhibit a small FE, which, similar to MAE, is linked to the tracking algorithms’ ability to produce outputs with minimal deviations from the expert trace.

However, none of the algorithms produced BSI values that were significantly different from the expert trace, except for U-Net segmentation applied to SQ images. Because BSI depends on the FR of both the DL and NDL, the U-Net results suggest that larger amplitude movements may hinder its ability to track fascia depth or that segmentation performance differed between limbs during the same exercise. This discrepancy may reflect subtle limb-specific segmentation differences during large-amplitude movements, where small absolute depth errors can propagate into amplified bilateral ratios. With respect to correlation, the variance-based algorithms produced traces most similar to the expert annotations. U-Net segmentation exhibited weaker correlations than the variance-based methods but stronger correlations than the intensity-based approaches, which showed poor shape fidelity. Additionally, performance differences were observed between movement tasks for both MAE and correlation, with SQ images generally resulting in worse performance than QA images. This likely reflects fundamental differences between the two movements: squats involve larger amplitude motion and require coordinated activation of multiple muscle groups, whereas quadriceps activations are isometric and primarily engage a single muscle group [44].

A notable strength of the variance-based tracking algorithms is their resilience to sources of noise and decorrelation in the signal over time [28]. This quality improves tracking of relative fascia movement; however, PCA-based methods presented in this study are fundamentally limited by the arbitrary scaling of principal components. Rescaling is required to map components to image depth and provide meaningful positional information. As a result, variance-based algorithms achieved the highest correlation, reflecting accurate shape tracking, but underperformed U-Net segmentation in MAE and most functional measures. Based on these properties, PCA-based tracking algorithms favor applications where the relative movement of fascia is more critical than the true depth of the fascia, such as relative fascia movement being used as a control signal to a device [8].

On the other hand, U-Net segmentation tracking is best suited for applications that require accurate depth information of the fascia. However, U-Net segmentation tracking is reliant upon a labeled dataset to train the algorithm, which researchers may not have access to for their given movement task [34]. The possibility of a U-Net segmentation model generalized to many movements by training on a large, multi-exercise dataset poses a potential solution to this challenge.

To assess whether the observed performance reflects an inherent limitation of the U-Net architecture rather than insufficient training data, we performed an ablation analysis varying the amount of training data used per movement type (see Supplementary Material, Fig. 11). Segmentation performance improved with increasing training data for quadriceps activation, but only modestly for squats, suggesting the influence of training-set size is task-dependent. This likely reflects the greater image variability and movement complexity of the squat condition rather than dataset size alone. Furthermore, while the U-net architecture itself does not appear to be a limiting factor of segmentation performance, there still may be other architectures which outperform it, which could be explored in future investigations.

Traditional muscle fascia tracking algorithms, such as MPI and MBTA, consistently exhibited weaker performance than the other tracking methods. In the past, MPI tracking and cross-correlation fascia tracking have been able to achieve modest performance, but each has experienced limitations. MPI tracking is prone to errors related to noise or changes in the strength of US signals from reflectors over time [29], [30]. Cross-correlation tracking is known to experience drift over time, resulting in degradation of tracking performance as the duration of the image increases [30]. MBTA attempts to mitigate these limitations by fusing the two methods’ outputs together [29], [30]. However, MBTA still faces limitations in both MPI and cross-correlation tracking, as indicated by our results. Jin et al., who introduced this fascia tracking method, were able to achieve high accuracy with the algorithm but were reliant upon implementing four US transducers at the same position with different angles and then selecting the best resultant image on which to perform their tracking [29]. Therefore, results from this and prior studies demonstrate that MBTA can be used to effectively extract fascia position, but image quality and characteristics may greatly degrade the performance of this algorithm.

Overall, the results from this study provide clear indications about the performance of the three types of tracking algorithms which were implemented: intensity-based, decomposition-based, and deep-learning-based. Intensity-based algorithms such as MPI tracking and MBTA are less resilient to poor image characteristics and therefore are limited to use-cases which are consistently able to produce clear images. This presents a problem for applications which are imaging movements with high speeds or complexity as these often degrade image quality. Decomposition-based algorithms are able to provide strong tracking of the relative motion of contractions but are limited in their ability to directly measure the depth of tissues, limiting their use cases as well. Deep-learning-based approaches provide strong tracking of the depth of tissues and modest tracking relative contraction pattern; however, these approaches struggle with the limitation of large training datasets being required for them to function properly. This type of dataset is non-trivial to acquire and requires a significant amount of expert labeling.

These findings have direct implications for wearable health monitoring and rehabilitation technologies that rely on ultrasound-derived muscle function metrics. In continuous or ambulatory monitoring scenarios, accurate estimation of fascia depth may be critical for tracking muscle activation magnitude, fatigue, or asymmetry over time, favoring neural network segmentation-based approaches when labeled data are available. In contrast, applications that prioritize consistent tracking of relative contraction patterns, such as providing real-time biofeedback during rehabilitation exercises or generating control signals for assistive or wearable devices, may benefit from variance-based methods that exhibit high correlation with expert traces while maintaining low computational cost. By clarifying these trade-offs, this work supports informed algorithm selection within wearable musculoskeletal sensing pipelines, rather than advocating a single tracking approach for all use cases.

### A. Clinical and Practical Applications

These findings offer practical guidance for selecting a fascia-tracking approach based on the intended use case. In clinical monitoring contexts where absolute fascia depth informs decisions, such as tracking muscle thickness changes during rehabilitation, U-Net segmentation offers the strongest accuracy, provided sufficient labeled training data exists for the target population and imaging system. In contrast, real-time biofeedback applications, where the priority is consistent tracking of relative contraction timing and pattern rather than absolute depth, are well served by PCA-based methods, which require no supervised training and are computationally inexpensive enough for onboard, real-time deployment on wearable hardware. This distinction gives developers of wearable MSK-US systems a concrete basis for algorithm selection depending on available data, computational budget, and the clinical or performance application, rather than defaulting to a single method across all use cases.

### B. Limitations

Several limitations should be acknowledged. First, ground truth annotations were provided by a single expert, which may introduce bias and influence the performance of supervised learning approaches; future studies should incorporate multiple annotators to improve reliability. We were able to gather ground truth annotations from a second expert for a small subset of squat images from the dataset and compared the performance of the algorithms to both of the expert traces for those images using MAE, Correlation, FR, and FE. These error metrics exhibited similar results for both experts’ ground truth annotations. These results are provided in the Supplementary Material. Second, this work evaluated only five tracking algorithms selected as they represent the most prominently used, distinct fascia tracking paradigms rather than an exhaustive survey of all permutations of these existing methods. While the algorithms used in this work are representative of these distinct paradigms there is the potential that other permutations may observe altered performance. Furthermore, other approaches, such as optical flow-based tracking, were excluded as they target a related but distinct measurement (muscle thickness change rather than fascia interface displacement) and are noted here as a direction for future comparative work. Third, the dataset was limited to two movement types and a single WUS system, which may restrict generalizability to other tasks, muscle groups, or imaging devices due to device-specific signal and image generation characteristics. Finally, computational efficiency was assessed on specific hardware configurations, and results may differ on lower-resource systems.

## CONCLUSION

This study systematically compared five fascia-tracking algorithms for wearable M-mode musculoskeletal ultrasound against expert-annotated ground truth. U-Net segmentation provided the most accurate estimates of absolute fascia depth and functional error, while PCA-based methods best preserved relative contraction patterns without requiring labeled training data. These findings clarify the trade-offs between tracking accuracy, robustness, and computational cost across algorithm types, offering practical guidance for algorithm selection in wearable neuromuscular monitoring applications. Future work should focus on automated region-of-interest definition and real-time implementation to support deployment of these algorithms in clinical and performance settings.

## Data Availability

All data produced in the present study are available upon reasonable request to the authors

## APPENDIX

### A. Preprocessing Techniques

To evaluate the performance of various preprocessing schemes, four techniques were implemented on each image. These preprocessing schemes include: no preprocessing, a thresholding of pixel values by a sigmoid function, reconstructing the original image after a non-negative matrix factorization (NNMF Recon), and a singular value filter (SVF) (Table IV).

Thresholding based on the sigmoid function preprocessed the M-mode images by applying all pixel values for an image as inputs to the following sigmoid function equation:

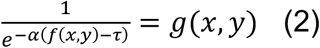

Where *f*(x,y) is the given pixel value, while α and τ are constants and equal to 0.05 and 127, respectively. This nonlinear scaling and thresholding of the images reduces the contrast at the brightest and darkest intensities while improving it within the midrange of pixel intensities.

To produce the NNMF reconstruction of images, each image resulting from the sigmoid threshold had an NNMF performed on it using five factors. This produced column vectors, spatial features related to the depth of interfaces in the image, and row vectors, time coefficients related to the presence of the spatial features over the time dimension. These spatial features and time coefficients can be multiplied to produce an approximation, or reconstruction, of the original image. For the purposes of this preprocessing approach, the number of factors was selected to be five to produce a reconstruction which contained only the most vital components of the original image and removed much of the speckle noise.

A SVF scales the singular values from a singular value decomposition before reconstructing the image [45]. This reduces or eliminates noise or irrelevant portions of the image as determined by a weighting function. The weighting function used for the SVF implemented in this work was:

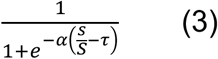

Where *s* is a singular value, *S* is the sum of all singular values, *α* is a constant equal to 40, and *τ* is a constant equal to 0.0275. This weighting function effectively reduces the contributions of information related to small singular values while maintaining information in the original image related to large singular values. An SVF using this weighting function was applied to each image with the sigmoid threshold applied beforehand.

### B. Impact of Preprocessing Techniques on Tracking Algorithms

The impact of preprocessing was assessed on MPI, MBTA, PCA, and CF-PCA tracking algorithms. All the metrics describing the performance of the tracking algorithms were calculated for each of the preprocessing techniques.

The MAE was unaffected by preprocessing for both the MPI and MBTA across both movement tasks. However, for the PCA and CF-PCA interface tracking, no preprocessing exhibited significantly smaller MAE than sigmoid thresholding (QA and SQ p<0.005) and in the case of SQ also NNMF reconstruction (PCA p<0.05; CF-PCA p<0.005) (Fig. 5).

**Fig. 4.**
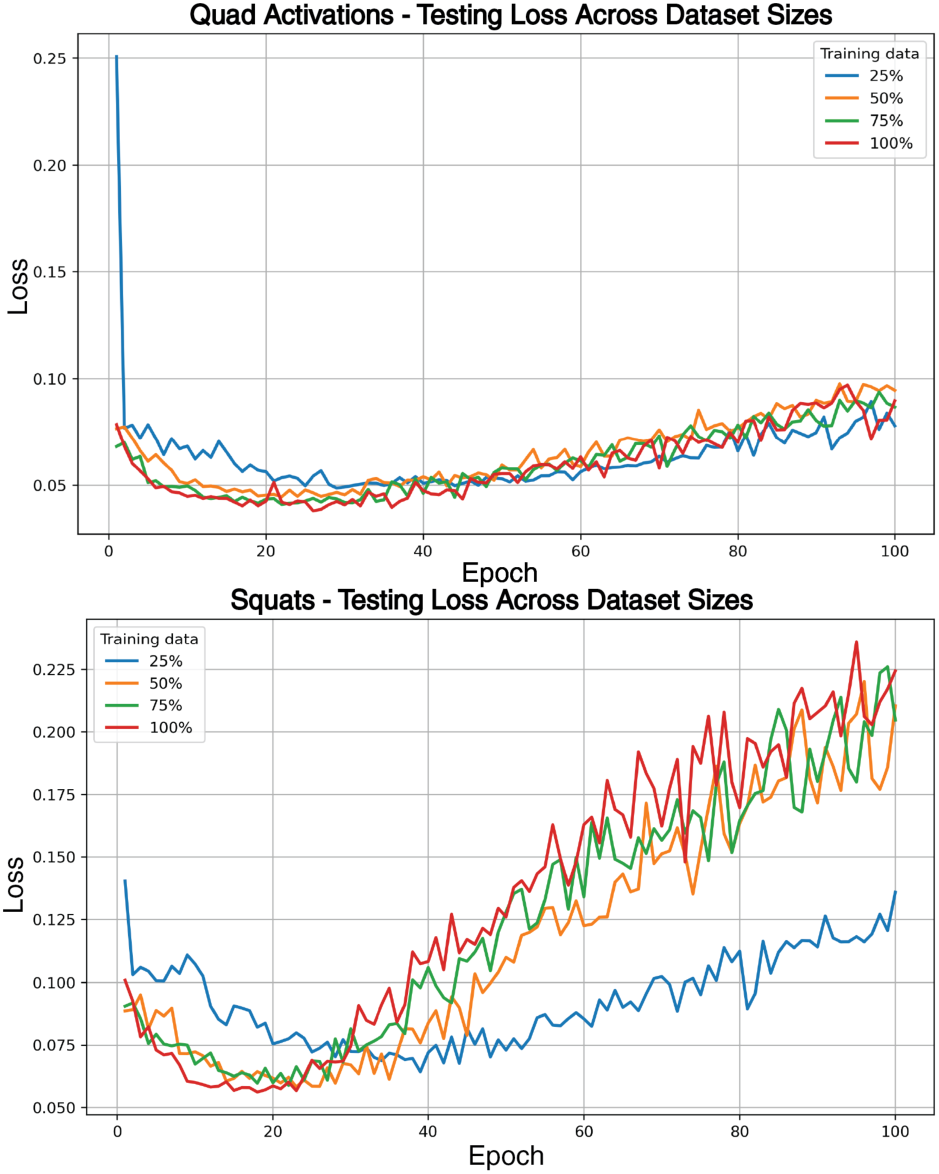
U-Net based testing (validation) data loss curves obtained using 25%, 50%, 75%, and 100% of the available training data from (top) quadriceps activation and (bottom) squat exercises.

**Fig. 5.**
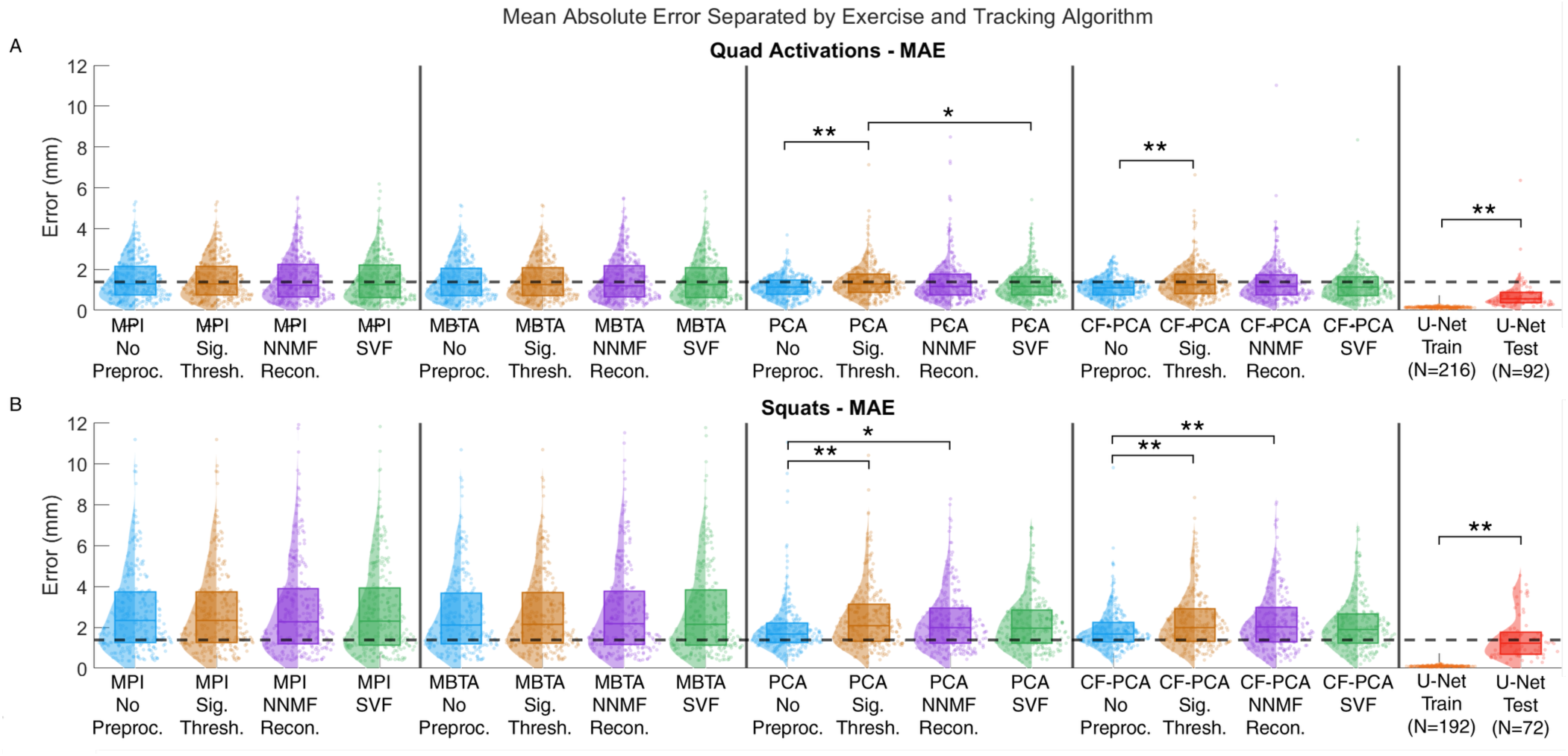
Distribution of mean absolute errors (MAE) for the combination of five tracking algorithms with four preprocessing techniques applied to (A) QA and (B) SQ. The MAE for each of the tracking algorithms were compared to the original, 10-pixel width (1.4 mm) of the expert traces, shown as the black dashed line. * *p*<0.05 and ** *p*<0.005.

Similarly, as in the MAE results, there were no significant differences between the preprocessing techniques for the correlation between the expert traces and the MPI and MBTA of both movement tasks (Fig. 6). For PCA and CF-PCA applied to QA, no preprocessing produced significantly higher correlations than sigmoid thresholding (p<0.005) and NNMF reconstruction (p<0.05). However, there were no significant differences between preprocessing techniques in correlations to the expert trace for PCA and CF-PCA tracking when applied to the SQ images.

**Fig. 6.**
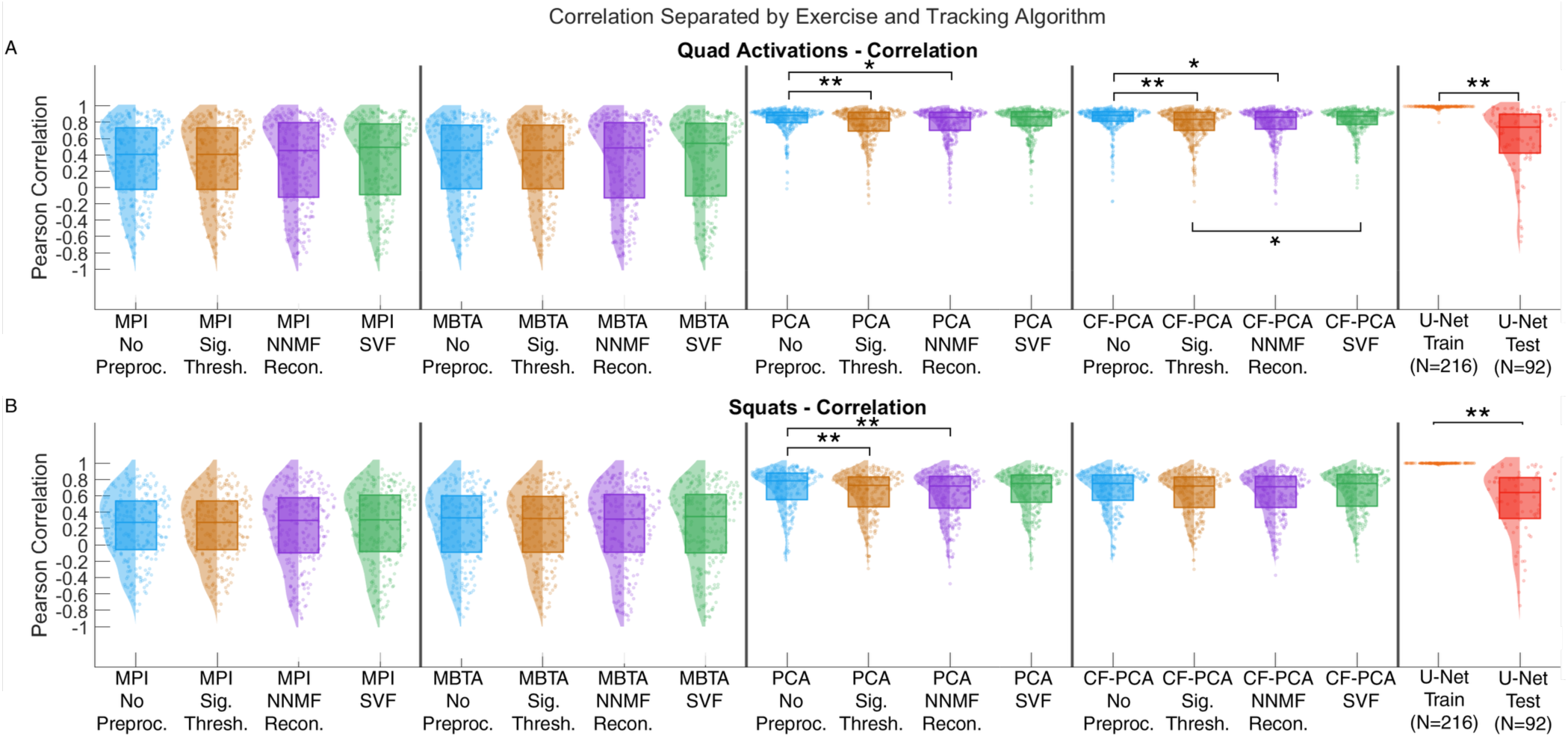
Distribution of Pearson’s correlation coefficients for the combination of five tracking algorithms with four preprocessing techniques applied to (A) QA and (B) SQ. * *p*<0.05 and ** *p*<0.005.

The FR extracted from the tracking algorithms was impacted by the preprocessing applied to the images (Fig. 7). Across all tracking algorithms, NNMF reconstruction and SVF resulted in significantly smaller FR in comparison to no preprocessing (QA and SQ p<0.005) and sigmoid thresholding, except for PCA and CF-PCA tracking applied to SQ images (QA and SQ p<0.005).

**Fig. 7.**
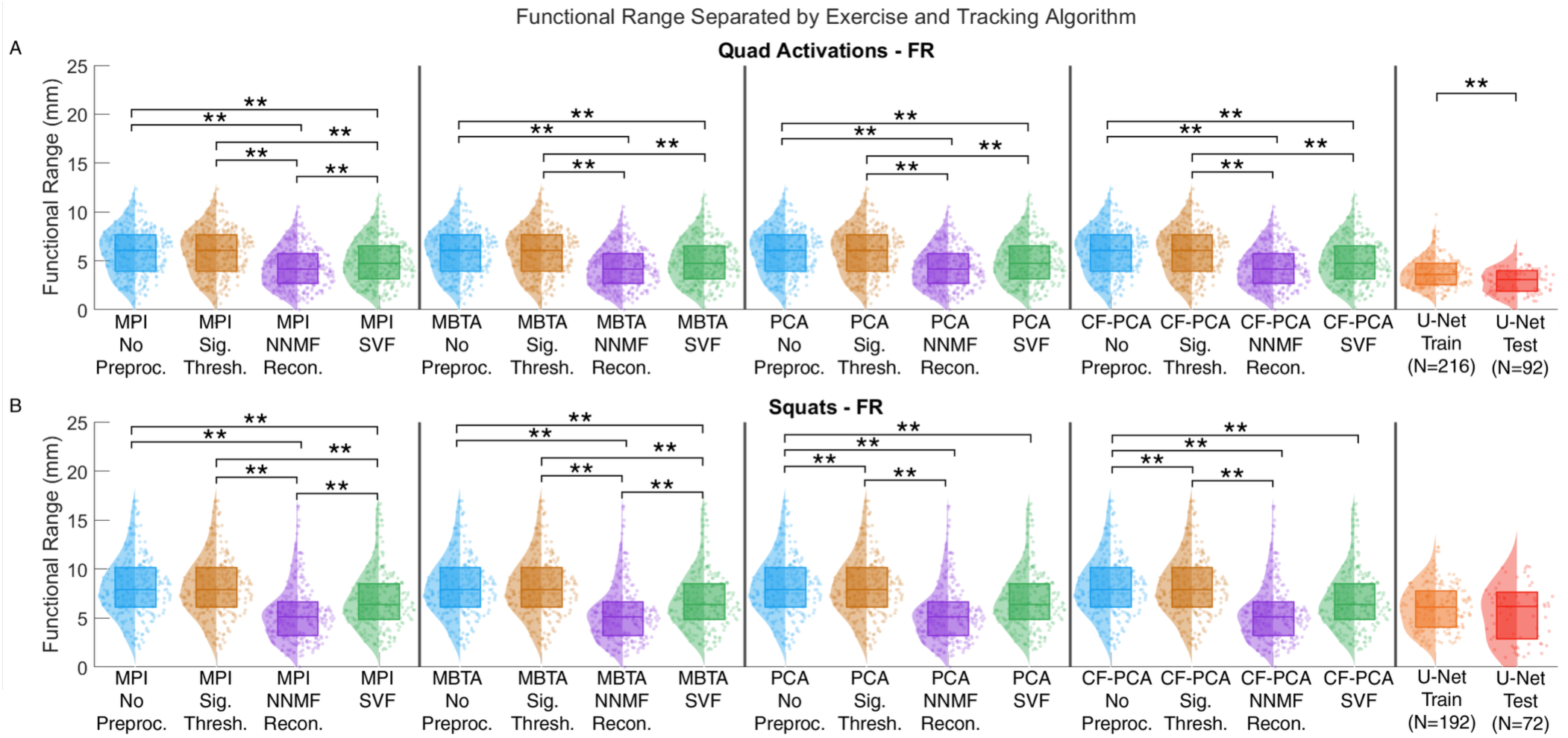
Distribution of functional ranges (FR) for the combination of five tracking algorithms with four preprocessing techniques applied to (A) QA and (B) SQ. * *p*<0.05 and ** *p*<0.005.

The FE was affected by the preprocessing, following the same relationships as seen in the FR (Fig. 8). The NNMF reconstruction had lower FE than no preprocessing or sigmoid thresholding (QA and SQ, p<0.005) across tracking algorithms. Furthermore, SVF had significantly smaller FE than no preprocessing (QA p<0.005; MBTA applied to SQ p<0.05; all other tracking algorithms applied to SQ p<0.005) apart from PCA and CF-PCA interface tracking for QA.

**Fig. 8.**
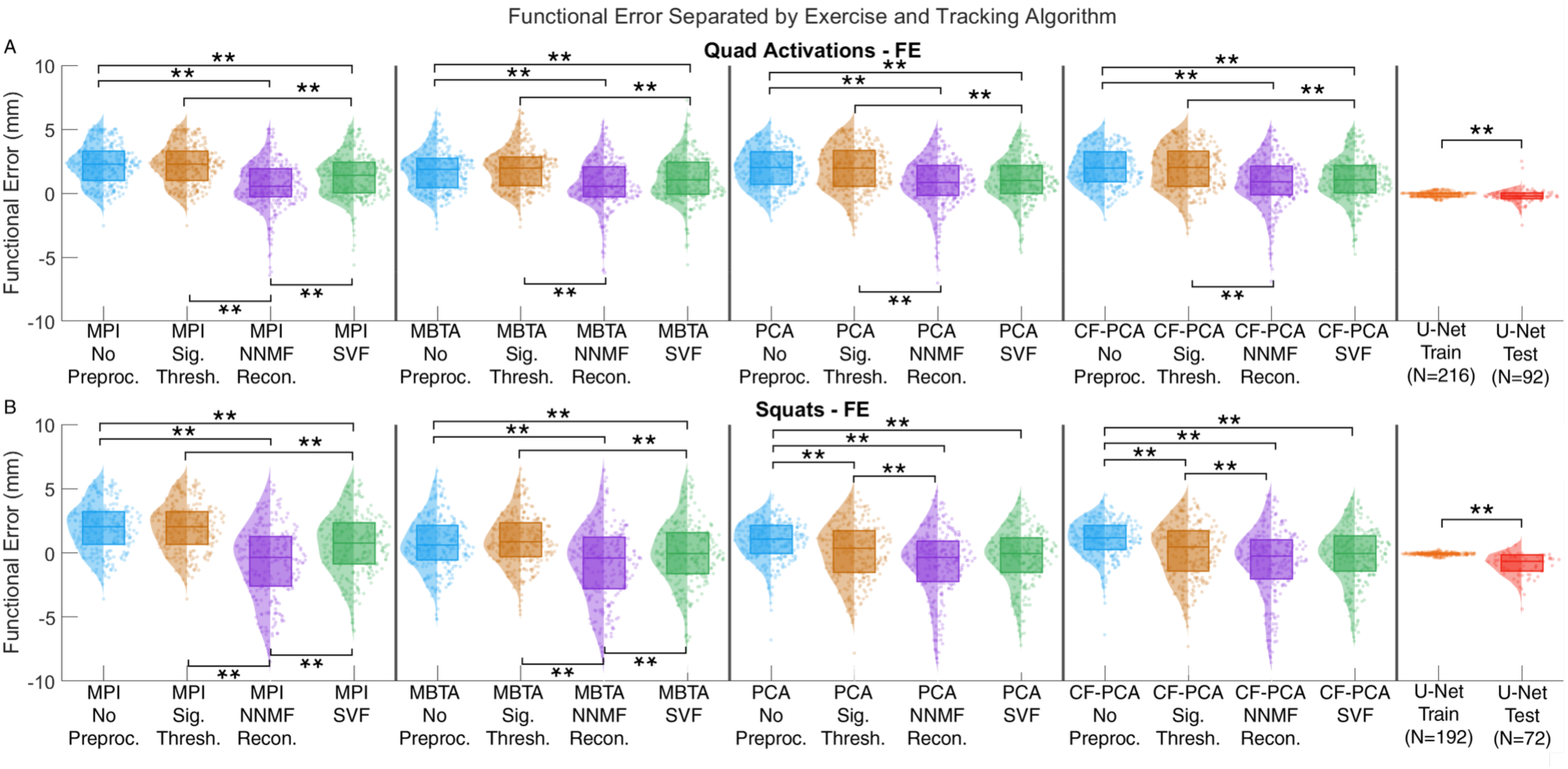
Distribution of functional error (FE) for the combination of five tracking algorithms with four preprocessing techniques applied to (A) QA and (B) SQ. * *p*<0.05 and ** *p*<0.005.

Preprocessing had an impact on whether the BSI calculated from the algorithms and expert traces were similar or not (Fig. 9). For QA, NNMF reconstruction (p<0.05) and SVF (p<0.005) lead to MPI and MBTA having significantly different BSI values from the expert trace. A similar effect was seen in the SQ data where nearly all tracking algorithms experienced different BSI values from the expert trace BSI for NNMF reconstruction (p<0.005) and SVF (MBTA and CF-PCA p<0.05; PCA p<0.005).

**Fig. 9.**
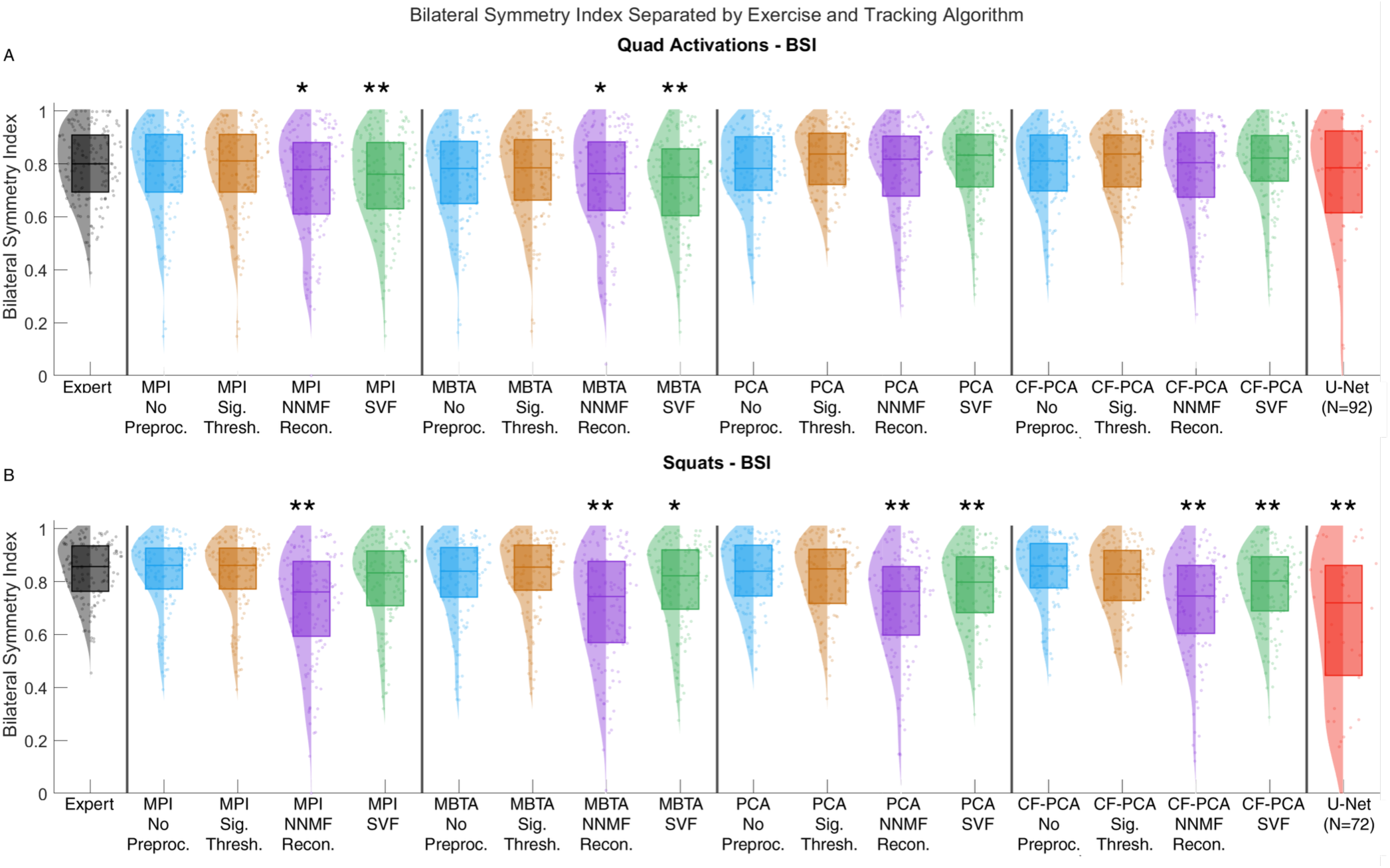
Distribution of bilateral symmetry indices (BSI) for the combination of five tracking algorithms with four preprocessing techniques applied to (A) QA and (B) SQ. * *p*<0.05 and ** *p*<0.005.

### C. Impact of Training-Set Size on U-Net Performance

To further investigate the influence of training-set size on the U-Net segmentation performance, an ablation analysis was performed using 25%, 50%, 75% and 100% of the available training images (per exercise) while maintaining the same testing data. For quadriceps activation, the segmentation performance improved progressively with increasing training data, with the Dice similarity coefficient (DSC) increasing from 0.6570 to 0.7250 between the 25% and 100% training conditions. In contrast, squat segmentation demonstrated only modest improvement, with DSC increasing from 0.4277 to 0.4320 across the same training conditions. Examination of the loss curves (Fig. 4) suggest that the influence of training-set size is task dependent. While the quadriceps activation (Fig. 4A) performance appears to benefit from the additional training data, the limited improvement observed during squatting (Fig. 4B) indicates that there are factors beyond the dataset size alone, such as increased image variability across subjects, and greater complexity of the movements which may have influenced the segmentation and consequentially tracking performance.

### D. Impact of Different Expert Traces on Tracking Algorithm Performance

A small comparison of tracking algorithm performance using the same image, but different expert annotated traces was performed in order to provide a preliminary indication of how tracking algorithms perform when compared to different expert traces. A second expert with 3 years of experience in musculoskeletal ultrasound image analysis and formal training in the WUS system’s image acquisition protocol provided annotations for a subset of 12 SQ images from the larger dataset. The resulting tracking algorithm outputs were then evaluated using MAE, correlation, FR, and FE. None of the distributions across all these error metrics and functional measures were found to be significantly different between the two reviewers using Kruskal-Wallis tests as described for the primary results (Fig. 10). While only a small percentage of total images from the larger dataset, this evaluation may indicate that tracking algorithm performance is relatively robust to the impact of different expert annotators.

**Fig. 10.**
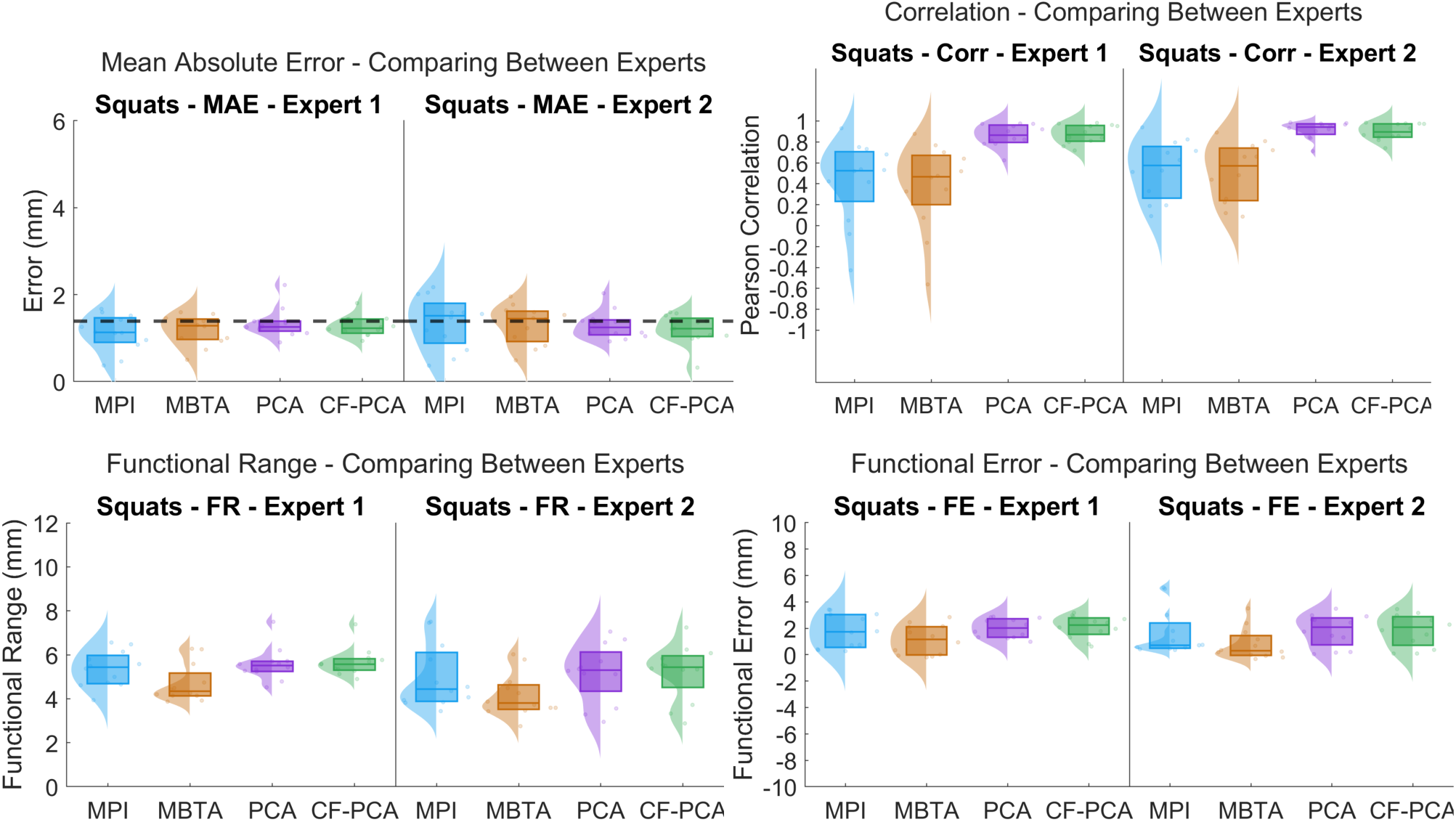
(Top Left) Distributions of mean absolute error for both expert annotators. (Top Right) Distributions for the correlation for both expert reviewers. (Bottom Left) Distributions for the functional range for both expert reviewers. (Bottom Right) Distributions for the functional error for both expert reviewers. No significant differences were found between the resulting distributions from each expert for any of these measures with *p*<0.05.

## Notes

### Competing Interest Statement

The authors have declared no competing interest.

### Author Declarations

IRB of George Mason University gave ethical approval for this work

### Funding

- Medical Technologies and Enterprise Consortium: W81XWH-15-9-0001, HT9425-24-9-0019

